# The impact of the one child policy on China’s infant mortality from 1970-1989: a quasi-experimental study

**DOI:** 10.1101/2022.03.09.22272156

**Authors:** Suman Chakrabarti, Esther Choo, Yao He

## Abstract

**Background:** China’s One Child Policy (OCP), enforced in 1980, was the largest fertility control policy in human history, and its effects are still widely debated. Previous analyses of the OCP have focused on population reduction and sex ratios, among others. This study evaluates the impact of the OCP on infant mortality in China using quasi-experimental methods.

**Methods:** Country-level panel data for China and 106 other countries were extracted from the World Bank data repository for the years 1970-1989. The primary outcome was infant mortality rate (IMR) defined as deaths among children <1 year of age per 1000 live births. The impact of the OCP was estimated using uncontrolled and controlled interrupted time series (ITS) designs that control for within group time varying characteristics. Models were further adjusted for a comprehensive set of covariates and corrected for first-order autocorrelation. Countries included in the control groups were identified using the method of synthetic controls and propensity score matching (PSM). Valid control groups reduced threats from selection bias and unobserved time invariant confounders.

**Results:** Adjusting for GDP per capita, GDP growth, women’s education, and population density, IMR decreased on average by 2.5 deaths per 1,000 annually before the OCP was enforced in China. The pre-OCP trends in IMR moved in tandem across China, synthetic and PSM controls, validating the parallel trends assumption. After the OCP, the trend in IMR in China increased by 2.24 (95% CI 1.39-3.09) per year, compared to the IMR trend in the pre-OCP period, comparing differences in China to synthetic controls. Results were similar and confirmed across PSM (1.96, 0.56-3.37) and uncontrolled ITS (2.14, 1.39-2.89) models. No significant immediate level change in IMR was observed in the controlled ITS models.

**Conclusions:** The reduction in IMR slowed down significantly after the OCP was enforced in China. The slowdown is likely due to a relative increase in the ratio of the IMR of females to males suggesting that practices such as female infanticide, abandonment, and neglect, stemming from a strong son preference were primary contributors. Coercive policies to reduce fertility can have unintended consequences.

## Introduction

In 1980, China started enforcing the one child policy (OCP), a nationwide fertility control measure and one of the largest natural experiments in human history^1^. Previous studies documented the impact of the OCP on demographic outcomes such as population control and sex ratio, but few articles focus on the OCP’s impact on infant mortality^1–4^. In the 1970s and early 1980s the most common causes of infant mortality in low- and middle-income countries were birth-related and malnutrition but it is unclear how the unique OCP affected infant mortality in China^5^. This study aims to evaluate the impact of the OCP on infant mortality rate (IMR) in China from 1970-1989.

China’s family planning efforts started in the early 1970s, but were not strictly enforced^6^. The OCP was proposed and adopted in 1979, with an initial strict implementation phase from 1980-1984^6,7^. After 1984 the OCP became the 1.5-child policy to accommodate rural traditions that favor boys so that rural families with a girl for the first pregnancy could have a second child^6^.

Around the same time the OCP was launched, China’s economic reform started in 1979, privatizing previously public social services^7^. The reform affected social determinants of child health such as healthcare and poverty throughout its decades-long implementation.

We hypothesize that the post-1980 trend in IMR in China is different from the pre-1980 trend, assuming that, in the absence of OCP, the pre-trend would have persisted after 1980 in China and the difference in the pre- and post-trend in China would have been the same as the difference in the control. This study will not only address gaps in the literature but also provide valuable insight on how similar fertility control measures could affect infant mortality.

## Methods

To address the research question, we use a controlled interrupted time series (ITS) analysis comparing China’s trend to a control group, between 1970-1989. The impact of China’s OCP was the difference-in-differences in the immediate level and trend change in IMR, comparing China and the control in this 20-year period.

### Data

The World Bank Development indicators provided annual country-level panel data for China and 106 other countries^8^. The outcome of interest was the IMR (deaths among infants <1 year per 1000 live births) from 1970 – 1989 for all countries. Other variables used in the analyses were total fertility rate (TFR), GDP per capita, adult female educational attainment, life expectancy at birth, and population density.

### Control group selection

The primary problem of causal inference is to estimate an appropriate counterfactual^9^. Given the data, a parsimonious model choice suggests that China’s own trend between 1970 and 1979 may be an appropriate counterfactual for what would have happened in the absence of the OCP^9^. However, history is a threat to internal validity in this uncontrolled ITS model because the OCP coincided with economic reform. We used the controlled ITS model to control for such within group time varying characteristics, selection bias, and unobserved time invariant confounding variables. We used two methods to identify our control groups: synthetic controls and propensity score matching (PSM).

The synthetic control method has been employed in a variety of social sciences including economics, public health, and politics^10^. This comparative case study method compares pre-intervention characteristics between potential control units and the intervention unit to knit together an optimal control similar in pre-intervention outcomes to the intervention unit^11,12^. Covariates used to identify the synthetic control were GDP per capita, TFR, women’s education, and population density (Figure A2). The final set of synthetic control countries are presented in Table 3 (N=80).

A second control group was selected through PSM which involved predicting countries with a high probability of having annual IMRs similar to those that China experienced between 1970 and 1979 (Figure A3)^13^. Covariates used to predict the PSM control group were life expectancy, TFR, female education, GDP per capita, and the pairwise interaction of these variables. The final set of PSM control countries are presented in Table 3 (N=540).

In a sensitivity analysis, an uncontrolled ITS model was presented using only China IMR data from 1970-1989^14^. The trend in IMR pre-OCP was compared with post-OCP level and trend.

### Statistical analysis

For the controlled ITS, we used a segmented linear regression model and compared pre-to post-OCP trends in the control to China^15^. The time variable was centered at 1979 (with 1977=- 2, 1978=-1,1979=0, 1980=1, and 1981=2, and so on) and the OCP dummy had a value of 1 for the years 1980-1989 and zero otherwise. For country *c* in year *t*,

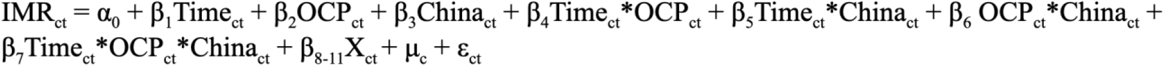

*α*_0_ estimates infant mortality in the control in 1979 with all covariates set to zero. *β*_5_ estimates the difference in pre-trend between China and control. *β*_6_ is the difference in level change in 1980 between China and control. *β*_7_ is the difference in slope change post-OCP between China and control. X_ct_ is a vector of covariates including log GDP per capita, women’s education, log population density, and GDP growth rate (Figure A1). μ_c_ is country random intercepts that account for country level heterogeneity. Errors are corrected for first-order autoregressive correlation^16^.

Synthetic and PSM controls were constructed in STATA version 14^17^. All analyses for the controlled and uncontrolled ITS were conducted in R version 4.0.0 with the *nlme* package (v3.1-148)^18,19^.

## Results

From 1970-1979, the mean TFR, mean life expectancy at birth, and mean years of women’s education were comparable between China and the two control groups (Table 1). China had lower mean IMR, lower mean GDP per capita, higher GDP growth rate, and larger overall population than the two control groups. Among the adjustment variables, the variations in GDP growth over time comparing post-to pre-OCP are different among China and the two controls; the other three variables had parallel upward trends from 1970-1989 (Figure A1).

**Table 1:** Summary statistics for China, Synthetic Control and Propensity Score Matched Control, 1970-1979.

|  | China |  | Synthetic control |  | PSM control |  |
| --- | --- | --- | --- | --- | --- | --- |
|  | Mean | SD | Mean | SD | Mean | SD |
| Infant mortality rate | 64.48 | 10.16 | 90.47 | 8.62 | 68.11 | 7.73 |
| Women's education | 3.17 | 0.45 | 3.17 | 0.33 | 4.61 | 0.45 |
| Total fertility rate | 4.13 | 1.05 | 4.20 | 0.30 | 5.27 | 0.37 |
| Life expectancy at birth | 63.22 | 2.44 | 58.35 | 1.59 | 61.80 | 1.42 |
| Population density | 96.07 | 5.37 | 122.64 | 7.18 | 63.01 | 3.95 |
| GDP per capita | 266.75 | 30.78 | 3546.30 | 342.28 | 2949.34 | 343.29 |
| GDP growth rate | 5.27 | 5.29 | 2.40 | 3.87 | 3.48 | 4.73 |
| Population, million | 902 | 50.4 | 166 | 11.1 | 14 | 1.1 |
| **N** | **10** |  | **40** |  | **270** |  |

Infant mortality rate per 1,000 live births, GDP per capita in constant \$, GDP per capita growth (annual %), Life expectancy at birth years, Total Fertility Rate (births per woman), Population density (people per sq. km), Women's education in single years (age 15-45 years). Synthetic controls include Cuba, India, Nepal, Portugal. Propensity Score Matched controls include Algeria, Botswana, Chile, Colombia, Costa Rica, Cuba, Dominican Republic, Ecuador, Fiji, Georgia, Guyana, Honduras, Jamaica, Kenya, Rep. Korea, Malaysia, Mexico, Nicaragua, Oman, Pakistan, Panama, Philippines, Sri Lanka, Suriname, Thailand, Tunisia, and Zimbabwe.

According to the adjusted ITS analysis using synthetic controls (Table 2), the annual decrease in mean IMR in China from 1970-1979 is 0.52 (95% CI -1.15, 0.1) per 1,000 live births higher than the annual decrease in an average control country (p = 0.10), suggesting that the pre-OCP trend in the synthetic controls is not significantly different from that in China. In 1980 the mean IMR in China is 1.00 (95% CI -2.87, 0.87) per 1,000 live births lower than the mean IMR for an average control country (p = 0.29). The difference in the annual change of mean IMR comparing post-to pre-OCP in China is 2.24 (95% CI 1.39, 3.09) per 1,000 live births higher than the difference comparing post-to pre-OCP in an average control country (p < 0.001; Figure 1).

**Table 2:** Estimates for the impact of the One Child Policy on infant mortality, 1970-1989.

|  | **Model 1** |  | **Model 2** |  | **Model 3** |  |
| --- | --- | --- | --- | --- | --- | --- |
|  | Uncontrolled ITS |  | ITS with Synthetic Controls |  | ITS with PSM Controls |  |
| $\beta_1$: Time | -2.53 | [-4.91, 0.16] | -2.31<sup>***</sup> | [-3.05, -1.57] | -2.06<sup>***</sup> | [-2.50, -1.63] |
| $\beta_2$: OCP | - | - | -0.50 | [-1.35, 0.36] | -0.51 | [-1.10, 0.08] |
| $\beta_3$: China | - | - | -48.26 | [-285.35, 188.82] | 31.03 | [-61.53, 123.59] |
| $\beta_4$: Time x OCP | - | - | 0.35 | [-0.04, 0.73] | 0.58<sup>***</sup> | [0.31, 0.86] |
| $\beta_5$: Time x China | - | - | -0.52 | [-1.15, 0.10] | -0.95 | [-1.94, 0.04] |
| **$\beta_6$: OCP x China** | -1.67<sup>*</sup> | [-2.97, -0.37] | -1.00 | [-2.87, 0.87] | -1.08 | [-4.21, -2.05] |
| **$\beta_7$: Time x OCP x China** | 2.14<sup>***</sup> | [1.39, 2.89] | 2.24<sup>***</sup> | [1.39, 3.09] | 1.96<sup>**</sup> | [0.56, 3.37] |
| $\beta_0$: Constant | 231.11 | [-352.21, 814.43] | 395.50<sup>***</sup> | [229.83, 561.16] | 166.49<sup>***</sup> | [124.35, 208.63] |
| **N** | **20** |  | **100** |  | **560** |  |

OCP: one child policy; PSM: propensity score matching; ITS: interrupted time series
All models are adjusted for log GDP per capita in constant US\$, women's education (aged 14-45) in years, log population density (persons/km square), growth rate of GDP.
Model 1: No control group.
Model 2: Control group is comprised of Cuba, India, Nepal, and Portugal. Model adjusted for country random intercepts.
Model 3: Control group is comprised of Algeria, Botswana, Chile, Colombia, Costa Rica, Cuba, Dominican Republic, Ecuador, Fiji, Georgia, Guyana, Honduras, Jamaica, Kenya, Rep. Korea, Malaysia, Mexico, Nicaragua, Oman, Pakistan, Panama, Philippines, Sri Lanka, Suriname, Thailand, Tunisia, and Zimbabwe. Model adjusted for country random intercepts.
95% confidence intervals in brackets with standard errors corrected for first order autocorrelation. \* $p < 0.05$ , \*\* $p < 0.01$ , \*\*\* $p < 0.001$

**Table 3:**
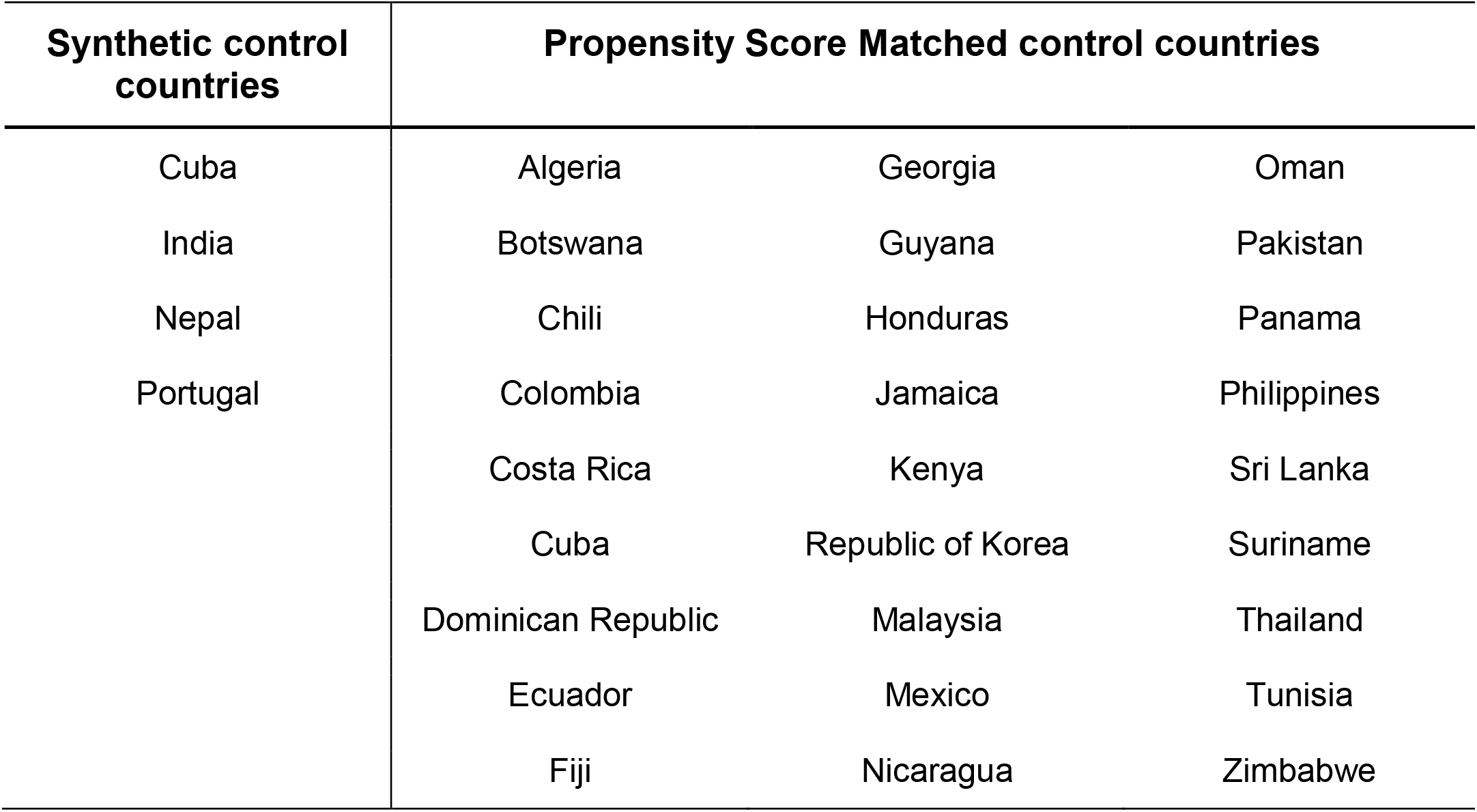
List of countries included in synthetic and propensity score matched controls.

| **Synthetic control countries** | **Propensity Score Matched control countries** |  |  |
| --- | --- | --- | --- |
| Cuba | Algeria | Georgia | Oman |
| India | Botswana | Guyana | Pakistan |
| Nepal | Chili | Honduras | Panama |
| Portugal | Colombia | Jamaica | Philippines |
|  | Costa Rica | Kenya | Sri Lanka |
|  | Cuba | Republic of Korea | Suriname |
|  | Dominican Republic | Malaysia | Thailand |
|  | Ecuador | Mexico | Tunisia |
|  | Fiji | Nicaragua | Zimbabwe |

**Figure 1:**
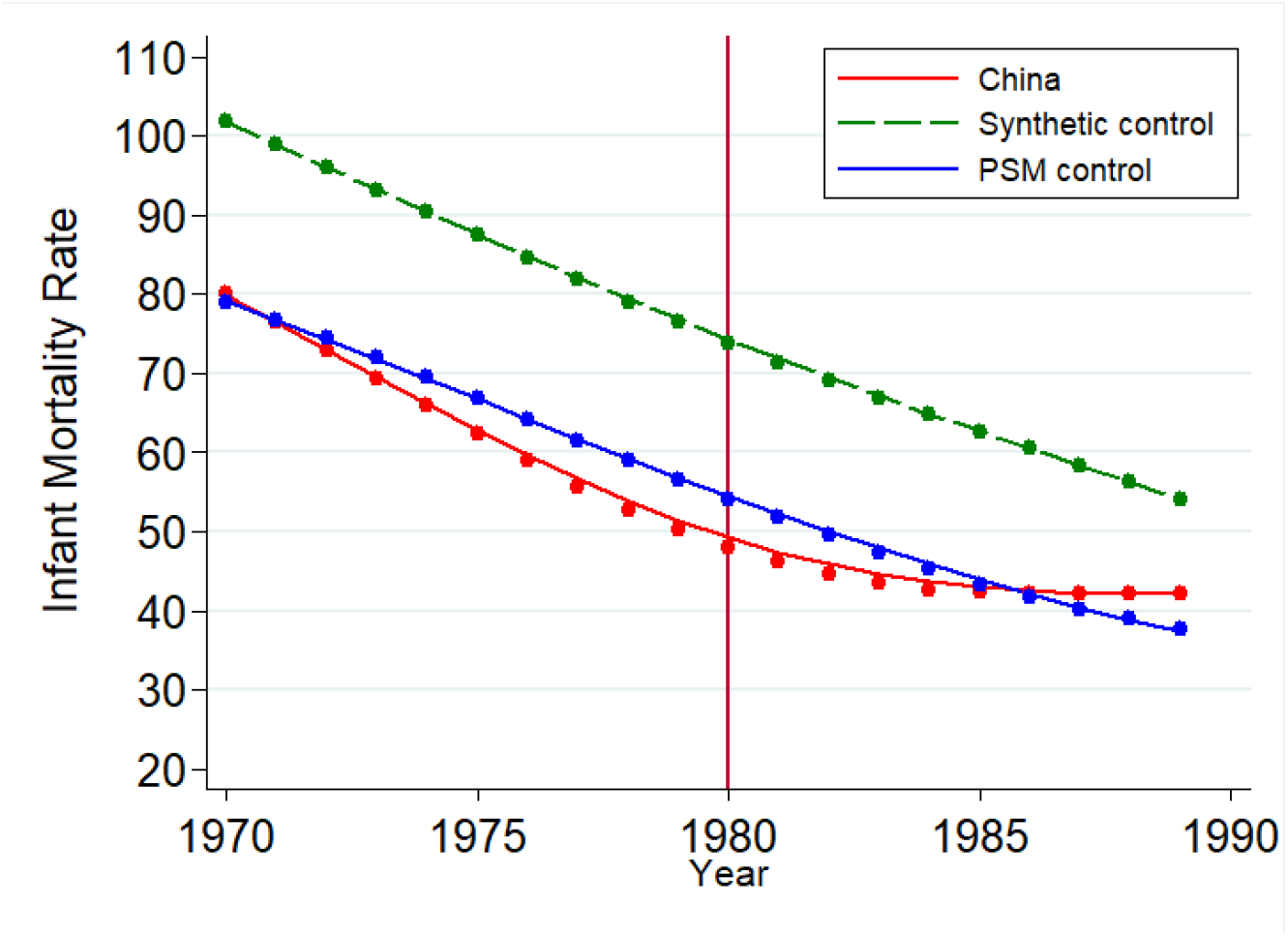
Trends in infant mortality across China, synthetic and propensity score matched controls, 1970-1989.

The findings of the adjusted uncontrolled ITS analysis and the adjusted ITS analysis using PSM controls are similar to the results reported above (Table 2). In the uncontrolled ITS analysis, the immediate decrease in IMR in China in 1980 is 1.67 (95% CI -2.97, -0.37) per 1,000 live births (p < 0.05). This immediate level change in 1980 occurs in other countries as demonstrated in the controlled ITS models. Thus, there was no immediate level change in China as a result of the OCP.

## Discussion

The IMR decreased at a slower pace for the first ten years post-OCP, a finding corroborated by all three models. This national policy to reduce fertility had negative spillover effects and unintended consequences resulting in higher than expected IMR. China’s progress on infant mortality may have been faster in the absence of this policy as modeled through the controls.

In terms of limitations, there is likely no perfect control available for China given its unique cultural heritage and political economy^20,21^. While the use of synthetic and PSM control groups with pre-OCP trends parallel to China’s minimizes selection bias, our estimates may have residual confounding due to differential trends in unmeasured time-varying covariates that change quickly overtime^22^. However, the similarity in the magnitude and range of estimates across all three models lends confidence that threats to internal validity for such biases are small.

## Conclusions

Globally, IMR is higher for males than females, while females have higher survival probability than boys^23^. Introduction of the OCP skewed the sex ratio towards males due to the greater preference for boys tied to future economic prospects and greater sense of support later in life^24^. Female IMR increased markedly in China after the policy due to sex-based discrimination at birth. This discrimination against girls led to abandonment, neglect of female infants and female infanticide^25^. This loss of female infants has been coined the “missing women” of China, a phenomenon that estimates 10-15 million women have gone missing as a result of China’s stringent fertility policies^26^.

This study demonstrates that coercive fertility control policies like the OCP can undermine progress on infant mortality reduction in the presence of son preference^27–34^. The findings are important for developing countries in the early stages of fertility transition and population control^35^. Investments in female education and non-coercive family planning programs to enhance access to contraception may succeed over top-down restrictions on fertility^36^.

## Data Availability

All data produced are available online at https://data.worldbank.org/

## Appendix 1

### Tables and Figures

**Figure A1:**
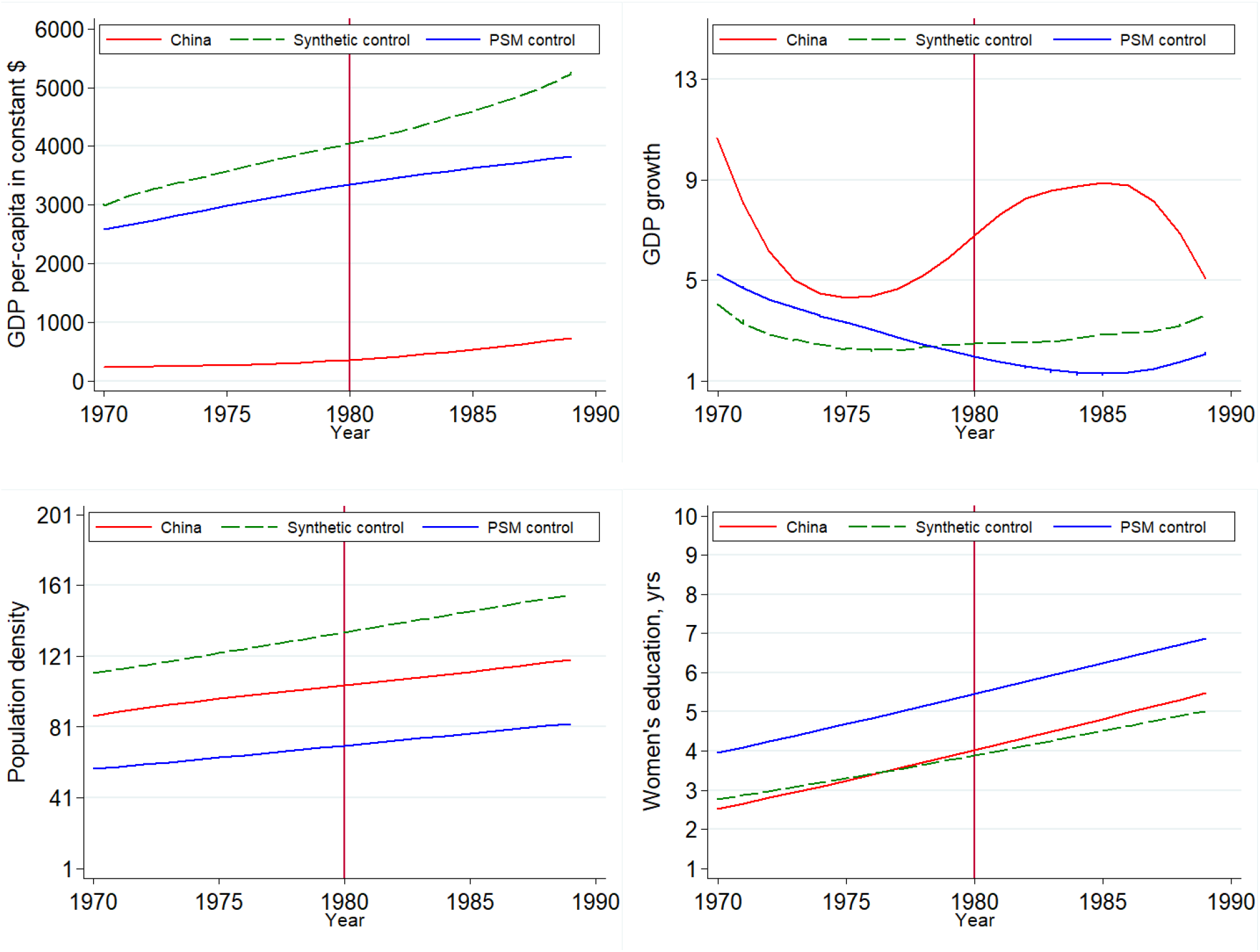
Trends in covariates across China, synthetic and propensity score matched controls, 1970-1989.

**Figure A2:**
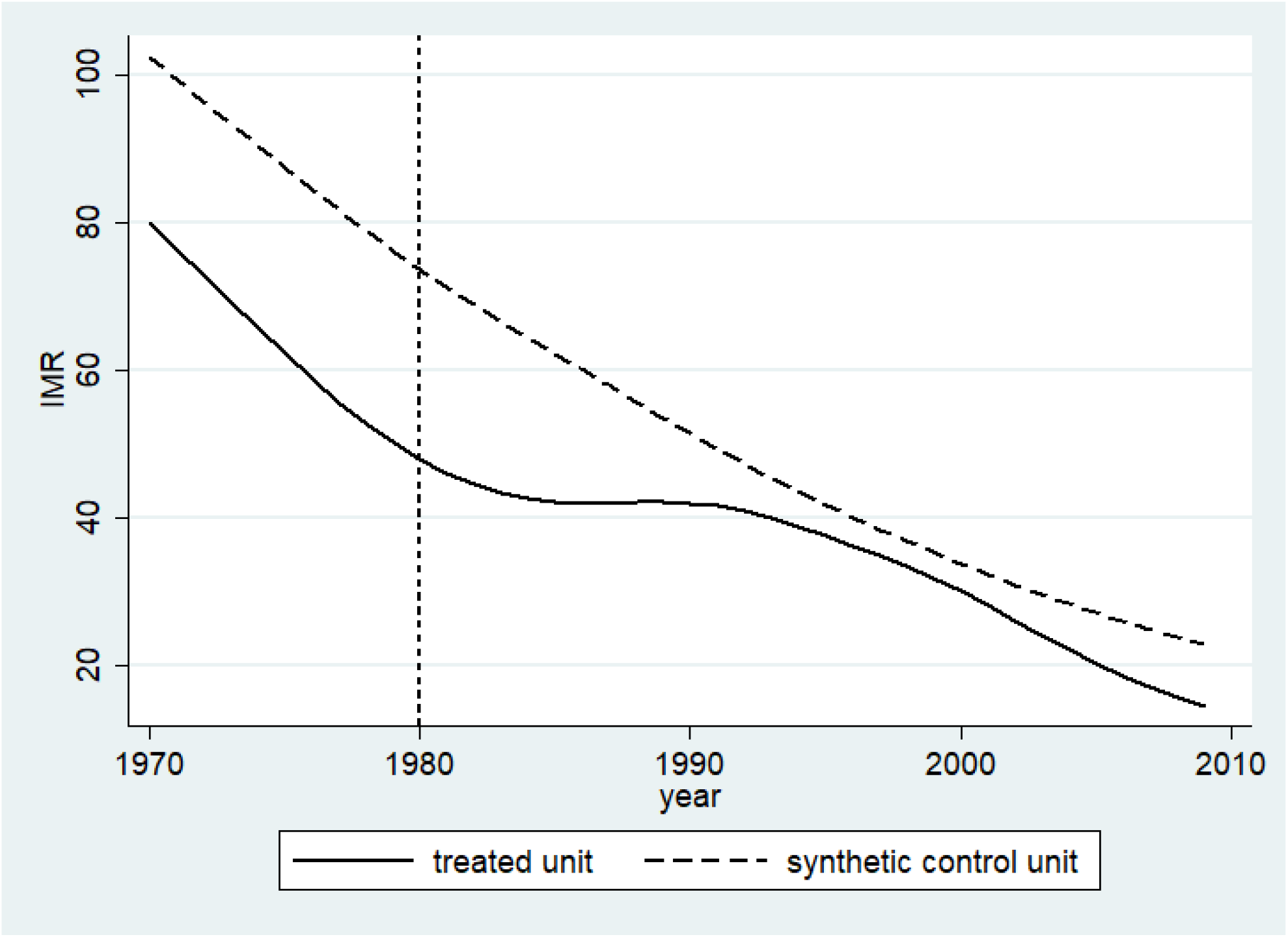
Synthetic controls weighted versus China, 1970-1989.

**Figure A3:**
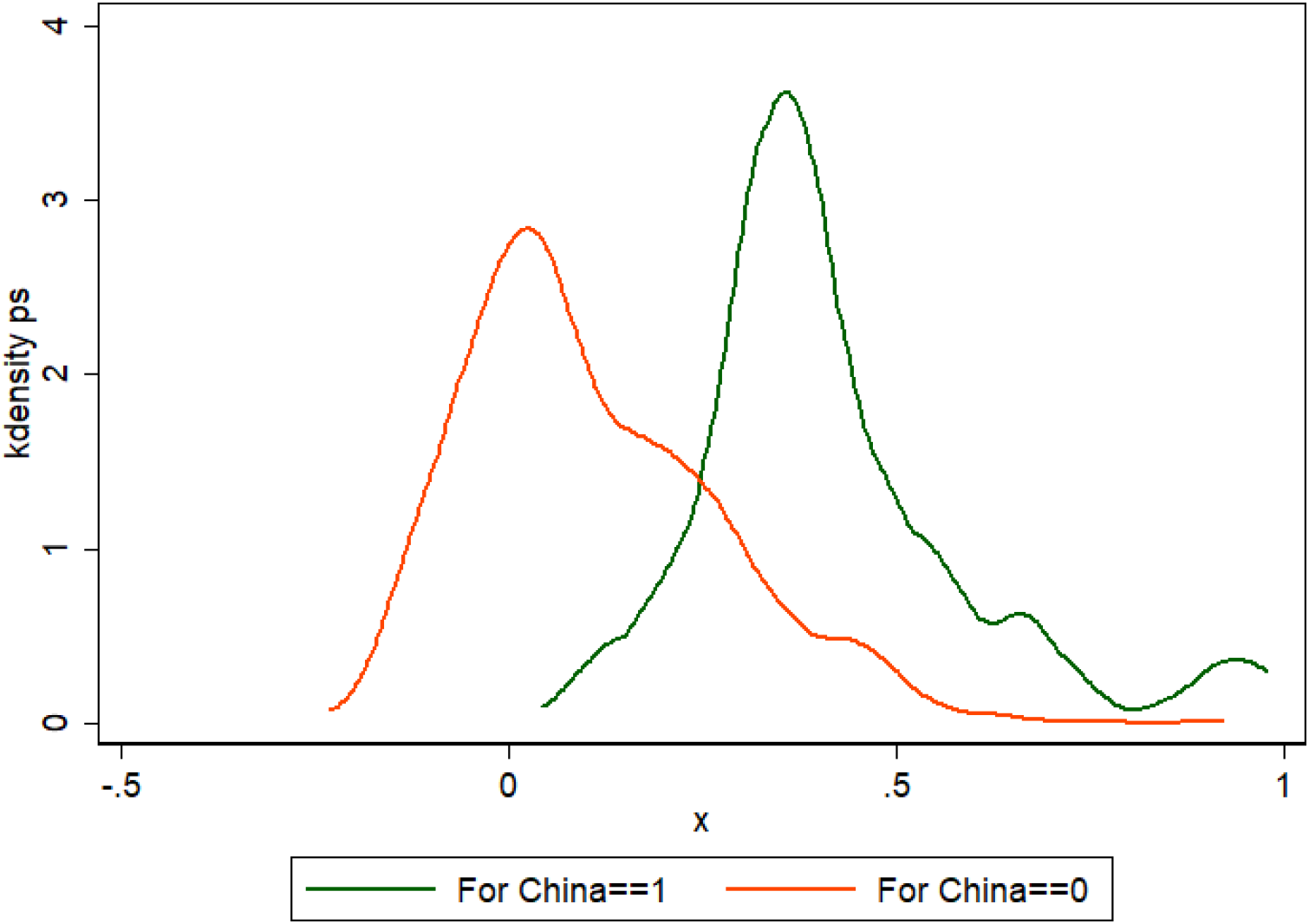
Common area of propensity score China versus other countries, 1970-1980.

## References

1. Hvistendahl, M. Has China outgrown the one-child policy? Science (2010) doi:10.1126/science.329.5998.1458.

2. Omran, A. R. The epidemiologic transition: A theory of the epidemiology of population change. Milbank Quarterly (2005) doi:10.1111/j.1468-0009.2005.00398.x.

3. Wang, F., Cai, Y., Shen, K. & Gietel-Basten, S. Is Demography Just a Numerical Exercise? Numbers, Politics, and Legacies of China’s One-Child Policy. Demography (2018) doi:10.1007/s13524-018-0658-7.

4. Goodkind, D. The Astonishing Population Averted by China’s Birth Restrictions: Estimates, Nightmares, and Reprogrammed Ambitions. Demography (2017) doi:10.1007/s13524-017-0595-x.

5. Ashworth, A. & Waterlow, J. C. Infant mortality in developing countries. Arch. Dis. Child. (1982) doi:10.1136/adc.57.11.882.

6. Zeng, Y. & Hesketh, T. The effects of China’s universal two-child policy. The Lancet (2016) doi:10.1016/S0140-6736(16)31405-2.

7. Hesketh, T., Lu, L. & Zhu, W. X. The effect of china’s one-child family policy after 25 years. New England Journal of Medicine (2005) doi:10.1056/NEJMhpr051833.

8. World Bank. World Development Indicators (WDI) | Data Catalog. Data Catalog, United Nations World Data bank (2019).

9. Varian, H. R. Causal inference in economics and marketing. Proc. Natl. Acad. Sci. 113, 7310–7315 (2016).

10. Gietel-Basten, S., Han, X. & Cheng, Y. Assessing the impact of the “one-child policy” in China: A synthetic control approach. PLoS One (2019) doi:10.1371/journal.pone.0220170.

11. Abadie, A. & Gardeazabal, J. The economic costs of conflict: A case study of the Basque country. Am. Econ. Rev. (2003) doi:10.1257/000282803321455188.

12. Abadie, A., Diamond, A. & Hainmueller, A. J. Synthetic control methods for comparative case studies: Estimating the effect of California’s Tobacco control program. J. Am. Stat. Assoc. (2010) doi:10.1198/jasa.2009.ap08746.

13. Dehejia, R. H. & Wahba, S. Propensity score-matching methods for nonexperimental causal studies. Review of Economics and Statistics (2002) doi:10.1162/003465302317331982.

14. Bernal, J. L., Cummins, S. & Gasparrini, A. Interrupted time series regression for the evaluation of public health interventions: A tutorial. Int. J. Epidemiol. (2017) doi:10.1093/ije/dyw098.

15. Shadish, W., Cook, T., Campbell, T. Experiments and generalized causal inference. Exp. quasiexperimental Des. Gen. causal inference (2005) doi:10.1198/jasa.2005.s22.

16. Wooldridge, J. M. Introductory Econometrics: A Modern Approach. Econ. Anal. (2003) doi:10.1198/jasa.2006.s154.

17. StataCorp. Stata Statistical Software:Release 14. Stata Statistical Software (2015).

18. Pinheiro, J., Bates, D., DebRoy, S. & Sarkar, D. R Core Team (2014). nlme: linear and nonlinear mixed effects models. R package version 3.1–117. URL http://cran.r-project.org/web/packages/nlme/index.html (2014).

19. Team, R. C. R: A Language and Environment for Statistical Computing. Vienna, Austria (2019).

20. The Economic Role of Political Institutions: Market-Preserving Federalism and Economic Development. J. Law, Econ. Organ. (1995) doi:10.1093/oxfordjournals.jleo.a036861.

21. Wu, X. Social change. in Handbook of Contemporary China (2011). doi:10.1142/9789814350099_0003.

22. Fewell, Z., Davey Smith, G. & Sterne, J. A. C. The impact of residual and unmeasured confounding in epidemiologic studies: A simulation study. Am. J. Epidemiol. (2007) doi:10.1093/aje/kwm165.

23. United Nations Inter-agency Group for Child Mortality Estimation (UN IGME). Levels & Trends in Child Mortality: Report 2019, Estimates developed by the United Nations Inter-agency Group for Child Mortality Estimation. United Nations Children’s Fund (2019) doi:10.1371/journal.pone.0144443.

24. Dewen, W. China’s urban and rural old age security system: Challenges and options. China World Econ. (2006) doi:10.1111/j.1749-124X.2006.00001.x.

25. Lai, D. Sex ratio at birth and infant mortality rate in China: An empirical study. Soc. Indic. Res. (2005) doi:10.1007/s11205-004-1542-y.

26. Shi, Y. & Kennedy, J. J. Delayed Registration and Identifying the Missing Girls in China. China Q. (2016) doi:10.1017/S0305741016001132.

27. Edlund, L. Son preference, sex ratios, and marriage patterns. J. Polit. Econ. (1999) doi:10.1086/250097.

28. Pande, R. & Malhotra, A. Son Preference and Daughter Neglect in India. International Centre of Research on Women (2006).

29. Kureishi, W. & Wakabayashi, M. Son preference in Japan. J. Popul. Econ. (2011) doi:10.1007/s00148-009-0282-3.

30. Haughton, J. & Haughton, D. Son preference in Vietnam. Stud. Fam. Plann. (1995) doi:10.2307/2138098.

31. Chung, W. & Das Gupta, M. The decline of son preference in South Korea: The roles of development and public policy. Popul. Dev. Rev. (2007) doi:10.1111/j.1728-4457.2007.00196.x.

32. Arnold, F., Choe, M. K. & Roy, T. K. Son preference, the family-building process and child mortality in india. Popul. Stud. (NY). (1998) doi:10.1080/0032472031000150486.

33. Leone, T., Matthews, Z. & Zuanna, G. D. Impact and determinants of sex preference in Nepal. Int. Fam. Plan. Perspect. (2003) doi:10.2307/3181060.

34. Obermeyer, C. M. & Cardenas, R. Son Preference and Differential Treatment in Morocco and Tunisia. Stud. Fam. Plann. (1997) doi:10.2307/2137891.

35. Bongaarts, J. & Casterline, J. Fertility Transition: Is sub-Saharan Africa Different? Popul. Dev. Rev. (2013) doi:10.1111/j.1728-4457.2013.00557.x.

36. Debpuur, C. et al. The impact of the Navrongo Project on contraceptive knowledge and use, reproductive preferences, and fertility. Stud. Fam. Plann. (2002) doi:10.1111/j.1728-4465.2002.00141.x.

